# One LLM is not Enough: Harnessing the Power of Ensemble Learning for Medical Question Answering

**DOI:** 10.1101/2023.12.21.23300380

**Authors:** Han Yang, Mingchen Li, Huixue Zhou, Yongkang Xiao, Qian Fang, Rui Zhang

**Affiliations:** Institute for Health Informatics, University of Minnesota, Minneapolis, Minnesota, USA; Division of Computational Health Sciences, Department of Surgery, University of Minnesota, Minneapolis, MN, USA; H. Milton Stewart School of Industrial & Systems Engineering, Georgia Institute of Technology, Atlanta, GA, USA

**Keywords:** Large Language Models, Ensemble Learning, Medical Question Answering, Healthcare AI

## Abstract

**Objective:** To enhance the accuracy and reliability of diverse medical question-answering (QA) tasks and investigate efficient approaches deploying the Large Language Models (LLM) technologies, We developed a novel ensemble learning pipeline by utilizing state-of-the-art LLMs, focusing on improving performance on diverse medical QA datasets.

**Materials and Methods:** Our study employs three medical QA datasets: PubMedQA, MedQA-USMLE, and MedMCQA, each presenting unique challenges in biomedical question-answering. The proposed LLM-Synergy framework, focusing exclusively on zero-shot cases using LLMs, incorporates two primary ensemble methods. The first is a Boosting-based weighted majority vote ensemble, where decision-making is expedited and refined by assigning variable weights to different LLMs through a boosting algorithm. The second method is Cluster-based Dynamic Model Selection, which dynamically selects the most suitable LLM votes for each query, based on the characteristics of question contexts, using a clustering approach.

**Results:** The Majority Weighted Vote and Dynamic Model Selection methods demonstrate superior performance compared to individual LLMs across three medical QA datasets. Specifically, the accuracies are 35.84%, 96.21%, and 37.26% for MedMCQA, PubMedQA, and MedQA-USMLE, respectively, with the Majority Weighted Vote. Correspondingly, the Dynamic Model Selection yields slightly higher accuracies of 38.01%, 96.36%, and 38.13%.

**Conclusion:** The LLM-Synergy framework with two ensemble methods, represents a significant advancement in leveraging LLMs for medical QA tasks and provides an innovative way of efficiently utilizing the development with LLM Technologies, customing for both existing and potentially future challenge tasks in biomedical and health informatics research.

## INTRODUCTION

Question Answering (QA) tasks in the medical domain involve a complex process of accurately interpreting and responding to healthcare-related queries[1]. QA tasks typically encompass two formats: open-ended and structured. In open-ended QA, respondents provide a complete sentence incorporating essential information in response to a question. In structured QA, the question is presented with several options, and the respondent selects the correct option or options by its corresponding identifier. Medical QA systems are designed to provide reliable and precise answers to questions ranging from disease symptoms and treatment options to medical research findings. These systems leverage advanced technologies like natural language processing (NLP) and machine learning approaches to understand and process medical terminology and concepts, making them invaluable tools for healthcare professionals and patients seeking medical information. The effectiveness of these systems is crucial, as they directly impact healthcare decision-making and patient care[2–5].

Previously, transformer models like BERT (Bidirectional Encoder Representations from Transformers) played a pivotal role in QA. For instance, He, Yun, et al.[6] infused disease knowledge into a basket of BERT-based models for health question answering, demonstrating the viability of disease knowledge infusion in NLP models. And Alzubi, Jafar A., et al.[7] developed another BERT-based model named CoBERT specifically designed for QA related COVID-19. The expansion of medical corpora and textual resources has necessitated leveraging these large datasets more effectively. This need has been met by the emergence of Large Language Models (LLMs) as a transformative approach to medical QA tasks. Pretrained on extensive and diverse datasets, LLMs like GPT-4 possess a deep understanding of language nuances and medical terminology, enabling them to generate highly accurate and relevant responses to medical queries[8]. They represent a significant milestone while dealing with natural language processing (NLP) tasks[9], such as text generation[10], question answering (QA)[11], Named Entity Recognition (NER)[12], etc. Moreover, LLMs include large-sized models like GPT-4 and Llama-2[13], as well as some relatively small yet efficient LLMs like Vicuna[14], and Stanford Alapaca. These LLMs, characterized by their vast size, have demonstrated remarkable capabilities in understanding and generating human language across a diverse array of domains[15].

Currently, the development and application of LLMs face distinct challenges based on their accessibility and capabilities[16]. On the one hand, closed-source models, such as GPT-3.5, GPT-4, and PaLM2[17], often maintained by large corporations, demonstrate advanced capabilities yet lack public accessibility. This limitation restricts their use in broader research and application contexts, including the situation of medical QA, especially when it comes to customized enhancements or applying potentially powerful techniques like fine-tuning[18]. On the other hand, open-source LLMs, like Open Llama2[19], Vicuna[14], and Alpaca[20], grant accessibility and transparency, as well as fine-tuning methods that are capable of making significant improvements[18]. A notable LLM instance in the medical QA context is PMC-LLama[20], exemplifying how fine-tuning an open-source LLM can lead to considerable improvements in its application, such as medical QA. However, this raises another significant challenge, which is the high cost associated with training and fine-tuning these models. The computational resources required for training LLMs with billions of parameters are substantial, making it impractical for individual researchers or small organizations to train these models from scratch[21]. In the healthcare and biomedical domain, the challenge is further compounded by the need for domain-specific knowledge[22–24]. Furthermore, selecting a suitable LLM for a specific biomedical application is not straightforward[25]. The choice of a model often depends on various factors, including the nature of the medical queries, the required level of accuracy, and the availability of domain-specific training data[10, 23, 26–27].

Ensemble method is a meta-approach stemming from machine learning techniques. In machine learning, ensemble involves combining multiple models to improve the overall performance, robustness, and reliability of predictions[28–29]. Techniques including voting[30–31], boosting[33], and bagging (bootstrap aggregating) [28–29, 32–33] are common ensembling approaches with better predictive performance by combining the predictions with multiple models or multiple past predictions. In the context of LLMs, model ensembles can potentially harness the diverse strengths of individual models to achieve superior results[31, 34–35]. These methods work effectively because they mitigate the weaknesses of single-model approaches. Each LLM, with its unique training data and architecture, may have specific strengths and biases[36–38]. Ensembling various models, each with distinct origins and structures, can equilibrate their inherent biases, mitigate overfitting, and enhance the generalization capacity for new data[39]. Importantly, this ensembling approach does not necessitate the models to be exclusively open-source or closed-source, nor does it demand extensive computational resources for its execution. Moreover, ensembling enables leveraging both domain-specific and general-purpose models. For instance, a general-purpose LLM might excel in understanding the context and semantics of a question, while a domain-specific LLM might provide more accurate technical information[20, 27, 40]. Combining these models can yield comprehensive and contextually relevant responses, crucial in medical scenarios.

Certain challenges lie in how to effectively ensemble LLMs, especially within the medical domain. This integration must consider factors such as the compatibility of different models, the method of aggregating their outputs and maintaining the interpretability of the responses. These considerations are crucial for ensuring that the ensemble not only performs well but also aligns with the stringent requirements of medical applications. There have been only a few studies diving into Ensembling LLMs to achieve a better prediction, like LLM-Blender[34], which implements PairRanker and Genfuser as an ensemble framework to generate consistently better responses for a given input. Similarly, the majority voting method proposed by Pitis, Silviu et al.[35] demonstrates potential. However, these studies primarily focus on open-ended tasks and do not delve into the specifics of medical QA, nor do they include domain-specific LLMs like PMCLlama2[21] or Medalpaca[28]. Furthermore, their applicability to structured QA with single-choice or multiple-choice questions in the medical domain remains unexplored, presenting unique challenges related to specificity, privacy, and data scarcity in medical contexts. This gap indicates the need for further research on the effective ensembling of LLMs tailored to the specific requirements of medical QA.

To address these limitations, we introduce LLM-Synergy, a novel ensembling framework tailored for medical QA, with two well-designed meta-learning ensembling methods, providing two innovative approaches combining the strengths of various LLMs, named Boosting-based Weighted Majority Vote Ensemble[35, 41–42] and Cluster-based Dynamic Model selection[43]. To validate the efficacy of LLM-Synergy and its ensemble methods, we conducted a case study using three medical QA datasets: MedMCQA[44], PubMedQA[45], and MedQA-USMLE[46].

Our contributions to this study include the following:

1. The development of innovative LLM ensemble methods, specifically the Boosting-based Weighted Majority Vote Ensemble and Cluster-based Dynamic Model Selection, offers new approaches, with zero-shot cases, in the medical QA field.
2. We implemented the ensembling methods for LLM methods and improved the performance by 5.98%, 1.09%, and 0.87% compared to the best-performing LLM on three medical QA datasets, the effectiveness of our ensemble methods. In each case, a tailored ensemble framework was created and adapted to the format of the QA dataset (single-choice or multiple-choice formats).
3. We conducted an error analysis to provide insights and directions for potential future enhancements in the field of medical QA, laying the groundwork for further improvements in this domain.

## METHODS

### Overview

The first step of our methods is investigating results with LLM-Synergy involves benchmarking leading LLMs including GPT-3.5-turbo, GPT-4, Llama2-13B, Vicuna-13B, Medalpaca-13B, PMC-Llama-13B, and a random guessing result as a reference. Within the benchmark, we conduct a sampled test, randomly drawing 200 QA pairs from the three medical QA datasets to assess the current capabilities of these LLMs in a medical context as a starting point. This benchmarking serves as a foundational analysis to understand the individual strengths and limitations of each LLM in handling medical QA tasks. Illustrated by Figure 1 overviewing of the whole pipeline of LLM-Synergy, following the benchmark assessment, the next phase focuses on the training process of our two proposed ensembling methods within LLM-Synergy: the Boosting-based Weighted Majority Vote Ensemble and the Cluster-based Dynamic Model Selection. The two approaches are designed to combine the unique capabilities of the selected LLMs, aiming to enhance the overall performance of medical QA systems. Moreover, the second method could be re-graded as an extensive version of the first one. By implementing these methods, we seek to address the shortcomings of relying on single models and reduce the need for extensive individual model training, thereby creating a more robust and efficient solution for medical QA.

**Figure 1.**
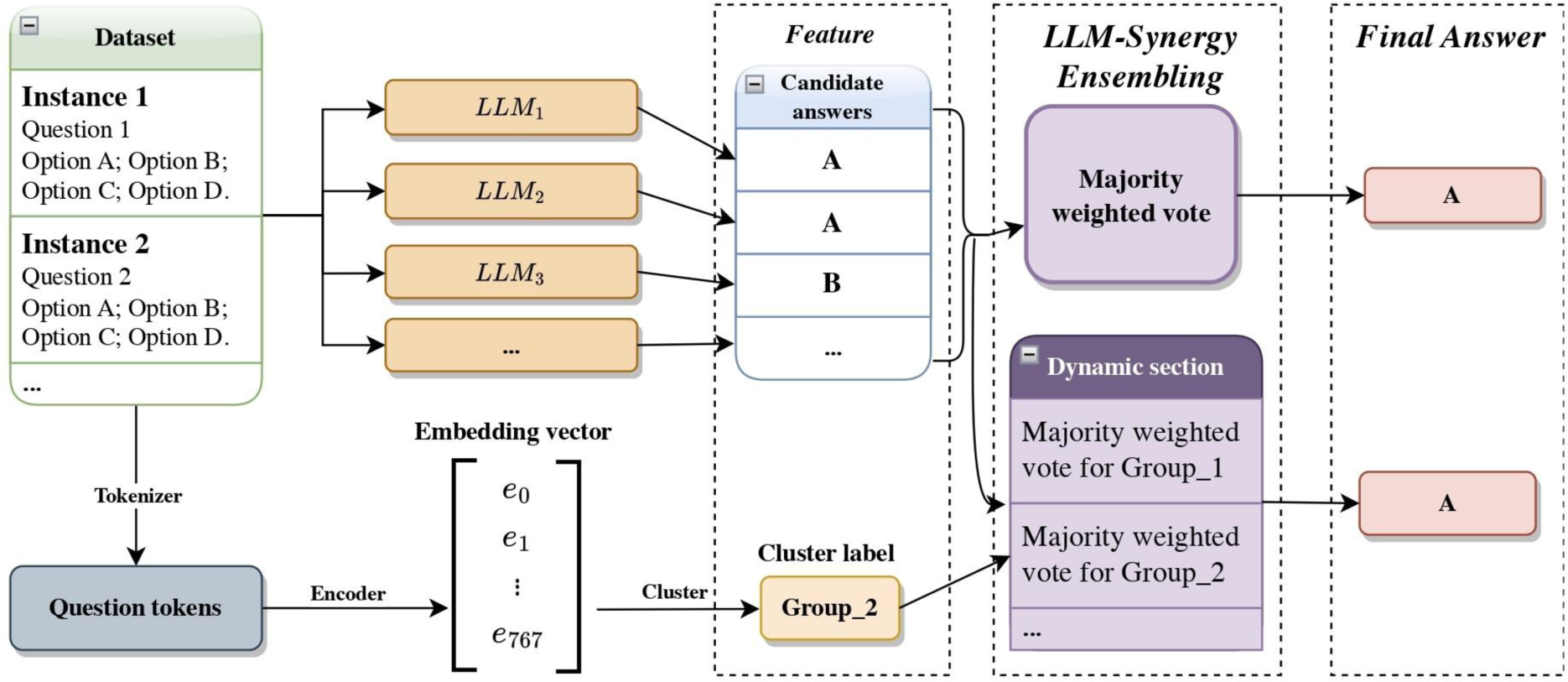
Overview of our LLM-Synergy Framework

### Dataset

We used three medical QA datasets for our model training and test: MedMCQA[44], PubMedQA[45], and MedQA-USMLE[46].

MedMCQA, released in March 2022 by Pal, Umapathi et al.[41], is a comprehensive multiple-choice question dataset derived from mock and past examinations of AIIMS and NEET-PG (Pal, Umapthi, et al. 2022), two prominent Indian medical entrance exams. It encompasses a training set with 182,822 questions and a test set comprising 4,183 questions, covering over 2,400 topics. Each question in this dataset presents four answer choices, labeled from A to D.

PubMedQA, introduced in September 2019 by Jin, Dhingra et al.[42], is a QA dataset curated from PubMed abstracts. It includes 1,000 questions reviewed by experts and 272,500 algorithmically generated QA pairs. The dataset’s primary task is to classify research questions into yes, no, or maybe answers, akin to multiple-choice questions. It is divided into three segments: PQA-L with 1k manually labeled pairs, PQA-U with 61.2k unlabeled pairs, and PQA-A featuring 211.3k artificially generated pairs. Here, we only implement a QA process without reasoning, which does not require a corresponding explanation of how the final answer is generated, which may lead to a relatively high accuracy score.

MedQA-USMLE, launched in September 2020 by Jin, Pan et al.[43], is an innovative dataset of multiple-choice questions tailored to the United States Medical Licensing Exams. This dataset encompasses questions in three languages: English, Simplified Chinese, and Traditional Chinese, with a total of 12,724, 34,251, and 14,123 questions in each respective language. Each question offers five choices, ranging from option A to E, sourced from professional medical board examinations. Here, we only experimented with the English QA parts.

The detailed LLM prompt can be found in the Appendix.

### LLM Benchmark on the three medical QA datasets

Ahead of implementing our ensembling framework, as a benchmark study, we evaluate the performance of various LLMs on 200 questions from each of the three QA datasets, respectively: PubMedQA, MedQA-USMLE, and MedMCQA, adhering to their specific answer formats. We evaluate our model’s performance against several robust baselines relying on the Language Model (LLM). Table 1 summarizes the six LLMs and one random guess predictor, along with the model characteristics including how many parameters within each LLM and a comprehensive description.

**Table 1:** The summarization of 7 LLMs running QA models.

| **QA Predictors** | **Model parameters** | **Description** |
| --- | --- | --- |
| GPT-4 | 1.76 trillion | GPT-4 is a substantial multimodal model designed to respond to questions by providing instructions fed to the GPT-4. |
| GPT-3.5-turbo | 20 billion | Same designed as GPT-4, however, GPT-3.5-turbo has fewer parameters than GPT-4. |
| Llama2-13B | 20 billion | Llama 2-13B is part of the Llama 2 series, representing a pretrained generative model. Tuned versions of Llama 2 utilize supervised fine-tuning (SFT) and reinforcement learning with human feedback (RLHF) to help generate the answers to given questions. |
| Vicuna-13B | 13 billion | Vicuna-13, similar in size to Llama2-13b, Vicuna is noted for its robustness and adaptability across different types of language processing tasks. |
| Medalpaca-13B | 13 billion | Medalpaca-13B is a substantial language model finely tuned for tasks in the medical domain. It stems from Llama (Large Language Model Meta AI) and boasts a significant parameter count of 13 billion. |
| MedLLama-13B | 13 billion | MedLLama-13B is initialized from LLaMA-13B and undergoes additional pretraining using a constructed medical corpus derived from 4.8M PubmedCentral papers and Medical Books. |
| PMC-LLama-13B | 13 billion | PMC-LLama-13B is the further tuned version of MedLLama-13B, with the pretrain and instruction-tuning methods. |
| Random Guess | 1 | A random guess predictor simply generates a random answer by the equal probability of each option, serving as a reference to compare the LLM-cased predictor. |

Figure 2 visualizes the performance of the selected predictor answering the three medical QA datasets. The benchmark graph illustrates that each predictor exhibits performance levels that exceed random guessing across various medical QA tasks, signaling the inherent capability of the LLMs to understand and process medical queries. Notably, different LLMs demonstrate particular strengths depending on the dataset; for instance, GPT-4 shows a marked proficiency in the MedMCQA tasks, while PMC-Llama-13B stands out in the PubMedQA context. These variations in model performance across tasks provide a solid foundation for the potential enhancement of accuracy through our subsequent ensemble work, suggesting that strategic combinations of these models could capitalize on their respective strengths and mitigate their individual weaknesses.

**Figure 2.**
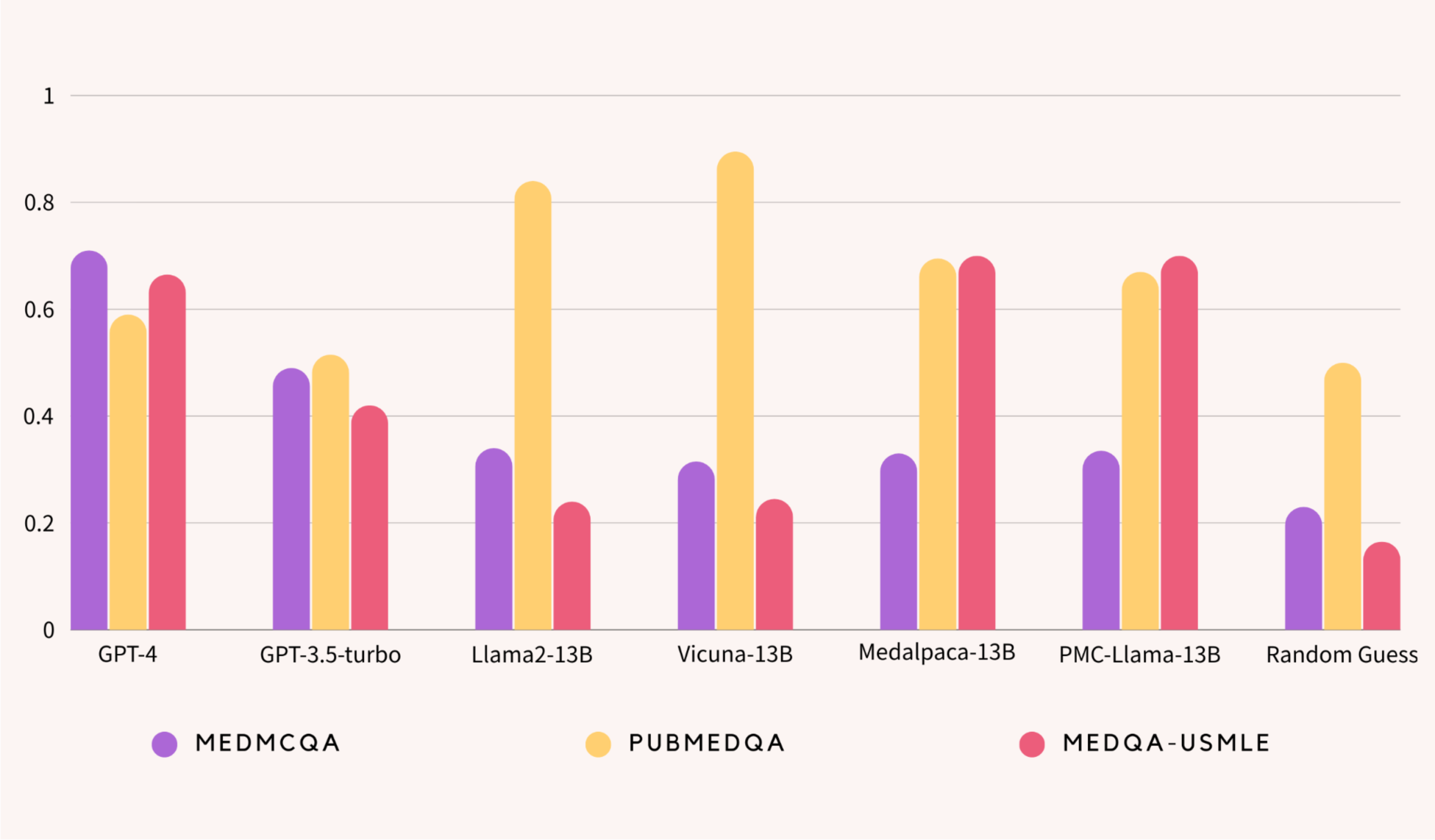
QA shows the accuracy of how each LLM performs on the three medical QA datasets.

Figure 3 shows the training process of Boosting-based Weighted Majority Vote Ensemble.

**Figure 3.**
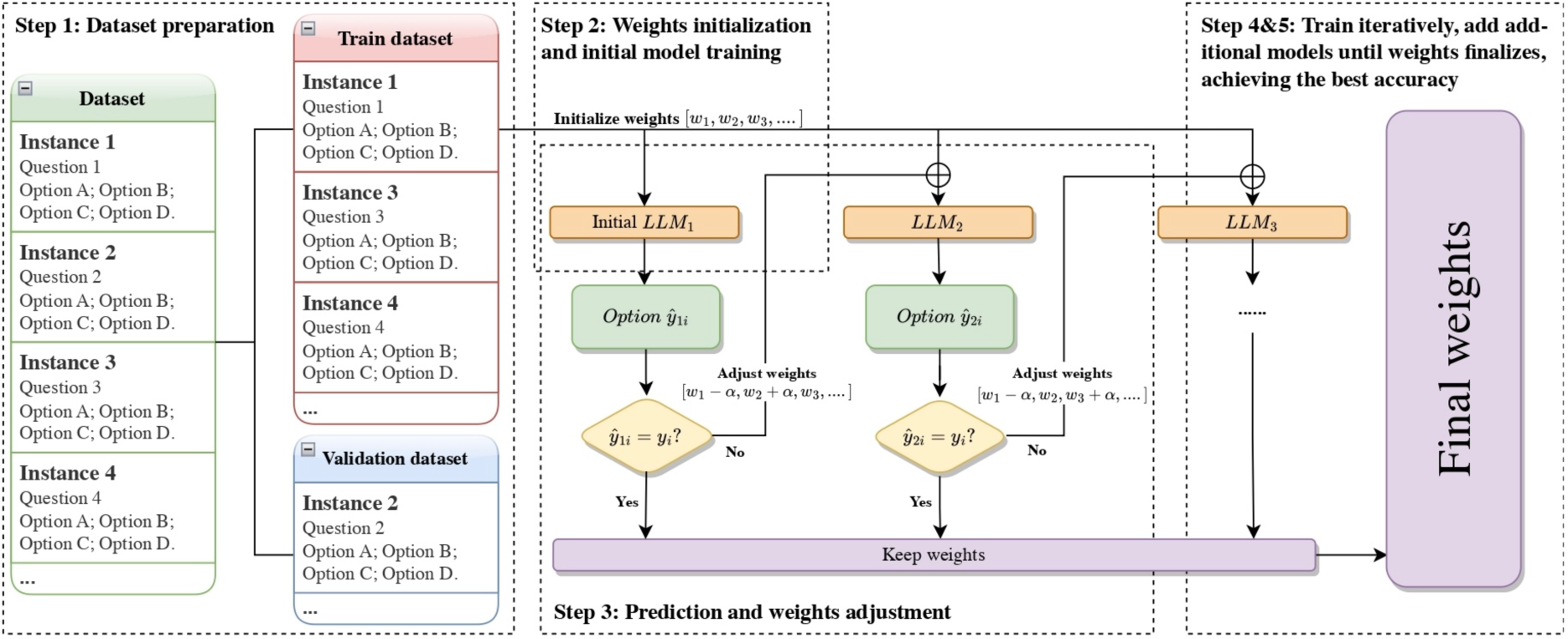
Training Process of Boosting-based Weighted Majority Vote Ensemble

#### Step 1: Dataset Preparation

For a given medical QA dataset, we split the dataset into a training set and a validation set, with a proportion of 80% and 20%. Each QA-pair instance in the dataset consists of a question and single-choice options.

#### Step 2: Weight Initialization and Initial Model Training

We assigned initial weights to all LLMs with weights (*w*_1_, *w*_2_,…) and initialized the starting status of ensembled LLMs with these weights. There may be different strategies for initialization, and we chose equally weighted initialization, which is the most common way of initialization[47].

#### Step 3: Prediction and Weight Adjustment

We chose a baseline LLM, denote as *LLM*_!_here, and focus on its wrong prediction: Use *LLM*_1_to predict answers for the training set and adjust the weights based on the prediction of instance: If the prediction, ŷ_1l_ for instance *i*, is incorrect, minus the weight of its corresponding LLM, *LLM*_1_, by an adjustment parameter *α*, i.e. *w*_i_ ← *w*_i_ + *α*, where *α* is a factor that increases the weight, indicating that the instance needs more attention in the next round of training.

#### Step 4: Iterative Training with Additional Models

For each subsequent LLM (*LLM*_2_, *LLM*_3_,…), repeat the prediction process. If a model correctly predicts the answer, we maintain the current weights. If the prediction ŷ_ni_ is incorrect, the weights were adjusted again.

#### Step 5: Finalize Weights of the Model Ensemble

After training and weight adjustments across all LLMs are completed, the final ensemble model is formed, which combines the individual LLMs with the final set of adjusted weights.

Figure 4 shows the training process of our second approach: Cluster-based Dynamic Model Selection, which serves as an extensive approach to the first one.

**Figure 4.**
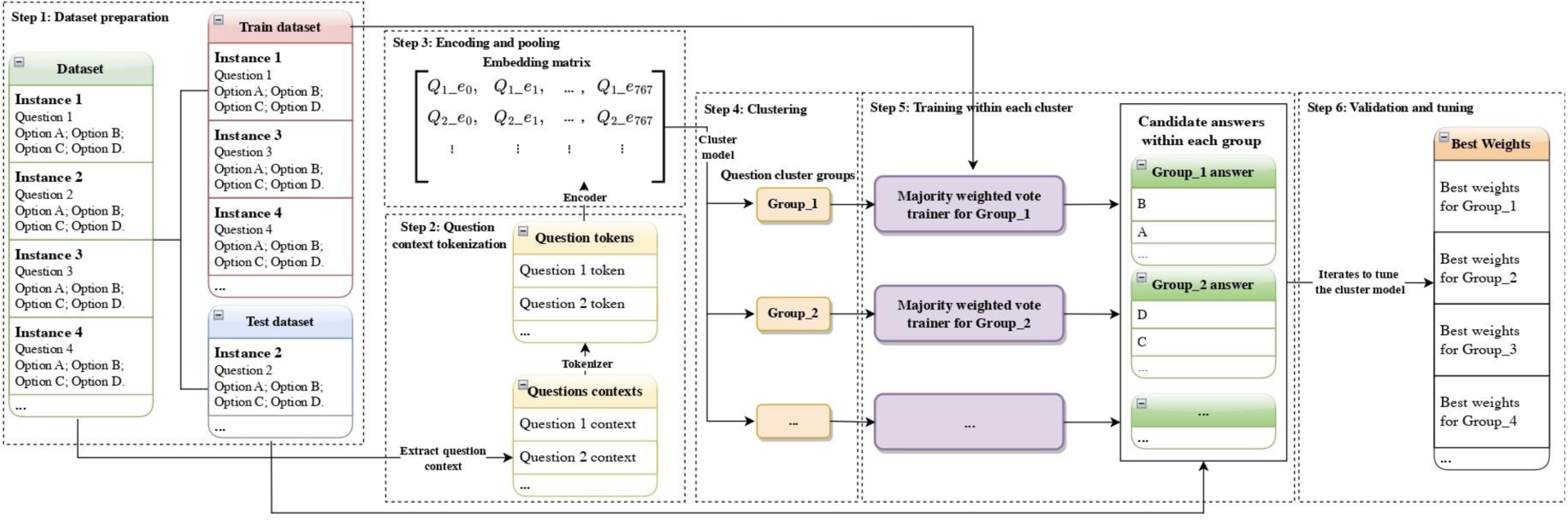
Training Process of Cluster-based Dynamic Model Selection Ensemble

#### Step 1: Dataset Preparation

Same as Part 1, for a given medical Question-Answering (QA) dataset, split it into a training set and a validation set with a ratio of 80:20. Each QA-pair instance in the dataset should consist of a question paired with single-choice options.

#### Step 2: Question Context Tokenization

Extract the context of each question from the QA pairs, and use the tokenizer, from Clinical-BERT in our case, to convert each question context into a series of tokens.

#### Step 3: Encoding and Pooling

Employ an encoder (same model as step 2, Clinical-BERT in our case) along with a mean-pooling method to encode each question’s tokens into a 768×1 embedding vector. Through this encoder part, we obtained a constructed embedding matrix for the entire set of questions, where each row represents the embedded vector of a question.

#### Step 4: Clustering

Apply a clustering model, KMeans in our experiment specifically, to the embedding matrix. This step assigns each question a cluster label, effectively reducing the dimensionality from 768 of an individual question vector to a single cluster group label being 1,2,3,…K. It serves as an unsupervised learning step that categorizes questions into different clusters based on their embeddings achieved after Step 3.

#### Step 5: Training Within Clusters

For each cluster group identified in the previous step, implement the Majority Weighted Vote trainer in Part 1, then determine the best combination of weights for the ensemble of LLMs within each group. These weights are adjusted to maximize the prediction accuracy of the ensemble within the specific context of each cluster.

#### Step 6: Validation and Tuning

Iteratively apply steps 2 to 5 to the validation set as well, and adjust and tune the hyperparameters of the clustering algorithm(K in KMeans in our case) to achieve the best overall accuracy on the validation set. This step is crucial for optimizing the model’s performance and ensuring that the clustering effectively captures the nuances of different question contexts.

### Evaluation

In the evaluation phase of our study, same as others[21, 36, 44, 46], accuracy was employed as the primary metric for assessment, given its congruence with the Micro F1 score in our specific experimental context of single-choice or multiple-choice QA tasks. Given the nature of multiple-choice QA datasets, metrics such as Recall and Precision are deemed inappropriate as they are sensitive to changes in the option numbering. For instance, adjusting the order of option labels may result in altering these metric values, when the number of options exceeds two. Based on these considerations, accuracy emerges as a more suitable evaluation metric in our case, providing a robust measure of overall correctness without being affected by variations in option numbering, which ensures a consistent and meaningful assessment of model performance in the specific context of our study.

The test set is distinguished from the training set used for model development and the validation set for hyperparameter optimization that we used within the ensemble training process. This distinct separation ensures an unbiased evaluation of the model’s true predictive capabilities. The test sets are derived from the subsets of the MedMCQA, PubMedQA, and MedQA-USMLE datasets, respectively. The MedMCQA test dataset consists of 4,183 QA pairs. The PubMedQA test dataset is even more extensive with 11,269 QA pairs. Whereas, the MedQA-USMLE dataset contains 1,272 QA pairs.

## RESULTS

In the presented study, the performance of various Large Language Models (LLMs) was evaluated against ensemble methods across three distinct medical QA datasets, MedMCQA, PubmedQA, and MedQA-USMLE. The performance metric, assumed to be an accuracy score, highlights the differential capabilities of each LLM and our two established predictors of ensemble methods. The detailed result can be seen in Table 2.

**Table 2.** Test set performance of each individual LLM and our ensemble approach.

| LLMs | MedMCQA | PubmedQA | MedQA-USMLE |
| --- | --- | --- | --- |
| Llama2-13B | 32.03 | 93.09 | 24.61 |
| Medllama-13B | 31.03 | 86.11 | 23.58 |
| Medalpaca-13B | 27.97 | 95.27 | 37.26 |
| Vicuna-13B | 26.13 | 93.15 | 24.14 |
| Majority Weighted Vote | 35.84 | 96.21 | 37.26 |
| Dynamic Model Selection | **38.01** | **96.36** | **38.13** |

For individual models, Llama2-13B demonstrated substantial proficiency in the PubmedQA dataset with an accuracy of 93.09%, indicating a strong alignment with the dataset’s characteristics. Conversely, its performance on the MedQA-USMLE dataset was considerably lower at 24.61%, suggesting a potential misalignment with this dataset’s attributes or a limitation in handling its complexity. Medllama-13B showed a similar trend, albeit with marginally lower accuracy figures across the board, peaking at 86.11% for the PubmedQA dataset.

Medalpaca-13B yielded a divergent performance profile, exhibiting a relatively lower accuracy of 27.97% on PubmedQA, while achieving the highest accuracy among individual LLMs on the MedQA-USMLE dataset at 37.26%. This suggests that Medalpaca-13B may possess particular strengths in processing the content typified within the MedQA-USMLE exam questions. Vicuna-13B, on the other hand, had the lowest accuracy scores for all datasets, which could indicate a general difficulty with the medical QA task as presented in these datasets.

The ensemble approaches, notably the Majority Weighted Vote ensemble and the Dynamic Selection ensemble, were developed to leverage the collective strengths of the individual LLMs. The Majority Weighted Vote Ensemble surpassed the individual model performances on the MedMCQA and MedQA-USMLE datasets and achieved a notable accuracy of 96.21% on PubmedQA. This enhancement suggests that a static weighted combination of model outputs can capitalize on the diverse expertise of each LLM to improve overall performance. Whereas, the Dynamic Selection Ensemble, which introduces a context-aware model selection strategy, further improved upon the Majority Weighted Vote ensemble’s performance, achieving the highest accuracy across all datasets: 38.01% on MedMCQA, 96.36% on PubmedQA, and 38.13% on MedQA-USMLE. The specific range of improvement varies from the variation of each LLM.

## DISCUSSION

The results underscore the variability in model performance across different medical QA contexts and the potential of ensemble methods to enhance prediction accuracy. The Dynamic Selection Ensemble, in particular, illustrates the advantages of a flexible approach that tailors model selection to the specific context of each question, aligning with the increasing demand for precision in medical informatics applications.

The LLM-Synergy ensemble framework is marked by its robust architecture, offering a suite of advantages that enhance its utility in the field of machine learning. Central to its design is the principle of scalability, which allows the framework to integrate an expanding roster of Large Language Models (LLMs) or extend to accommodate larger datasets with minimal structural adjustments. Flexibility is another cornerstone of the LLM-Synergy framework, demonstrated by its support for both zero-shot and multiple-shot applications, and its agnostic approach to model sourcing, accepting LLMs irrespective of their open-source or closed-source status. This versatility ensures that the framework is not limited by the availability or proprietary nature of models, thus broadening its applicability. Beyond its flexibility, LLM-Synergy is characterized by domain adaptability, with an underlying methodology that transcends the medical question-answering domain for which it was initially designed. Its principles are equally relevant to question-answering tasks in various other fields, indicating its potential for extensive adoption beyond the confines of healthcare. In terms of computational efficiency, LLM-Synergy stands out by harnessing the strengths of pre-existing, pre-trained models, thereby circumventing the necessity for extensive retraining. This attribute not only conserves computational resources but also expedites the deployment process. Additionally, the framework’s operational efficiency is exemplified by its modest demand for computing power, distinguishing it from many contemporary models that often require substantial computational investment. Another key advantage is the robustness of the ensemble approach, which integrates insights from multiple models to produce a more reliable and error-resistant output. By aggregating diverse perspectives, LLM-Synergy mitigates the influence of individual model biases or inaccuracies, resulting in a more dependable collective judgment.

Collectively, these advantages position the LLM-Synergy framework as a scalable, flexible, and computationally prudent choice for diverse machine learning applications, promising significant advancements in the realm of AI-driven problem-solving.

### Error Analysis

In the context of our LLM-Synergy framework, the efficacy of both ensemble methods is contingent upon the variance in performance among the incorporated LLMs. Within the MedMCQA dataset, the discrepancy in performance metrics among the LLMs was relatively narrow, with Llama2-13B achieving the apex at 32.03% and Vicuna-13B at the nadir at 26.13% in the individual case. Subsequent to integration into the ensemble framework, the LLMs demonstrated a capacity for reciprocal augmentation of their respective predictive strengths, culminating in an appreciable performance enhancement of 5.98%. In contrast, the PubMedQA dataset exhibited a more homogenized performance distribution, with even the least accurate LLM, Medllama-13B, attaining an accuracy rate of 86.11%. This scenario yielded an incremental, albeit less substantial, improvement post-ensemble due to the already elevated baseline accuracies. The MedQA-USMLE dataset, in other words, presents a distinctive case; Medalpaca-13B’s performance markedly surpasses its counterparts. In such instances, a static Majority Weighted Vote approach inherently biases the ensemble towards this single model, potentially allocating 100% of the weight to Medalpaca-13B and negating the contributions of other LLMs. Conversely, the Dynamic Model Selection method—owing to its context-sensitive algorithm—offers a degree of rectification by adjusting the ensemble’s reliance on the most apt LLM for a given question context.

### Limitation

The two frameworks within LLM-Synergy, Weighted Majority Vote Ensemble, and Cluster-based Dynamic Model selection, while robust and versatile, are not without limitations. One notable constraint is the dependency on the quality and diversity of the LLMs incorporated into the ensemble. The performance of the ensemble is inherently tied to the individual capabilities of the included models. If these models share common blind spots or biases, or if they are not sufficiently diverse in their approaches to problem-solving, the ensemble may not significantly outperform its individual constituents.

For the Weighted Majority Vote Ensemble, a key limitation lies in its static nature. The weights assigned to each model are fixed after training and do not adapt to the nuances of individual questions or contexts within the test set. This could lead to suboptimal performance in situations where the most appropriate model might change depending on specific question characteristics. The Dynamic Model Selection framework, while more adaptable, relies heavily on the clustering algorithm’s ability to meaningfully categorize questions. If the clustering does not effectively capture the relevant features that dictate which LLM would perform best, the dynamic selection process may not yield the intended improvements. Additionally, this method’s success is contingent upon the availability of a sufficiently large and representative validation set to fine-tune the model selection process. Moreover, both frameworks could still potentially be limited by computational constraints in practice, even if not computationally hungry as methods like fine-tuning or pretraining. Despite being designed to be resource-efficient, the process of integrating multiple complex models, especially when scaling up to include additional LLMs or accommodating larger datasets, could still demand significant computational resources.

Lastly, the frameworks assume that the best-performing LLMs on the validation set will continue to be the best choices for the test set. This may not hold if there are substantial differences between the validation and test sets, leading to a discrepancy between expected and actual performance. Therefore, while the LLM-Synergy frameworks offer promising approaches to ensemble learning in NLP, they must be applied judiciously, with an awareness of these potential limitations.

## CONCLUSION

The LLM-Synergy framework, with its boosting-based Weighted Majority Vote and cluster-based Dynamic Model Selection methods, represents a significant advancement in leveraging LLMs for medical QA tasks and provides an innovative way of efficiently utilizing the development with LLM Technologies, customing for both existing and potentially future challenge tasks in biomedical and health informatics research. Its ability to amalgamate the strengths of multiple models has demonstrated superior accuracy and robustness over individual LLMs. While the framework showcases scalability, flexibility, and adaptability across domains, it also presents opportunities for future enhancements, including increased model diversity, dynamic weighting, and broader domain applications. As such, LLM-Synergy not only addresses current challenges in natural language processing but also sets the stage for continued innovation in AI-driven problem-solving.

## DATA AVAILABILITY

The LLM-synergy framework used in this study is made publicly available at https://github.com-/serendipitYang/LLM-Synergy.

## CONTRIBUTORSHIP STATEMENT

HY served as the principal author, designing the entire experiment, writing experimental code, and the full manuscript. ML and YX collaborated on the data collection and analysis and also reviewed the paper. HZ assisted in conceiving the study design and reviewed the manuscript. RZ was responsible for auditing the feasibility of the research topic and refining the paper. QF created the graphical and workflow diagrams within the manuscript. All authors contributed significantly to the production and proofing of the manuscript.

## ACKNOWLEDGEMENTS

We would also like to acknowledge to the staff at BPIC of the University of Minnesota and the programmers from Dr. Rui Zhang’s group for their technical support.

## APPENDIX

Prompt for MedMCQA dataset:

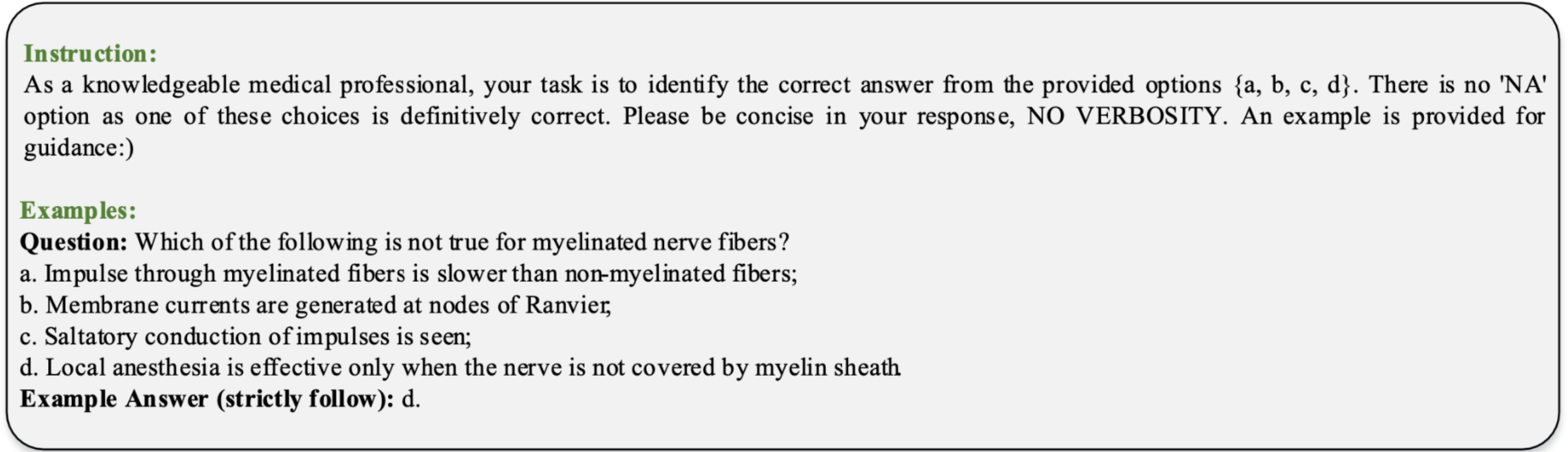

Prompt for PubMedQA dataset:

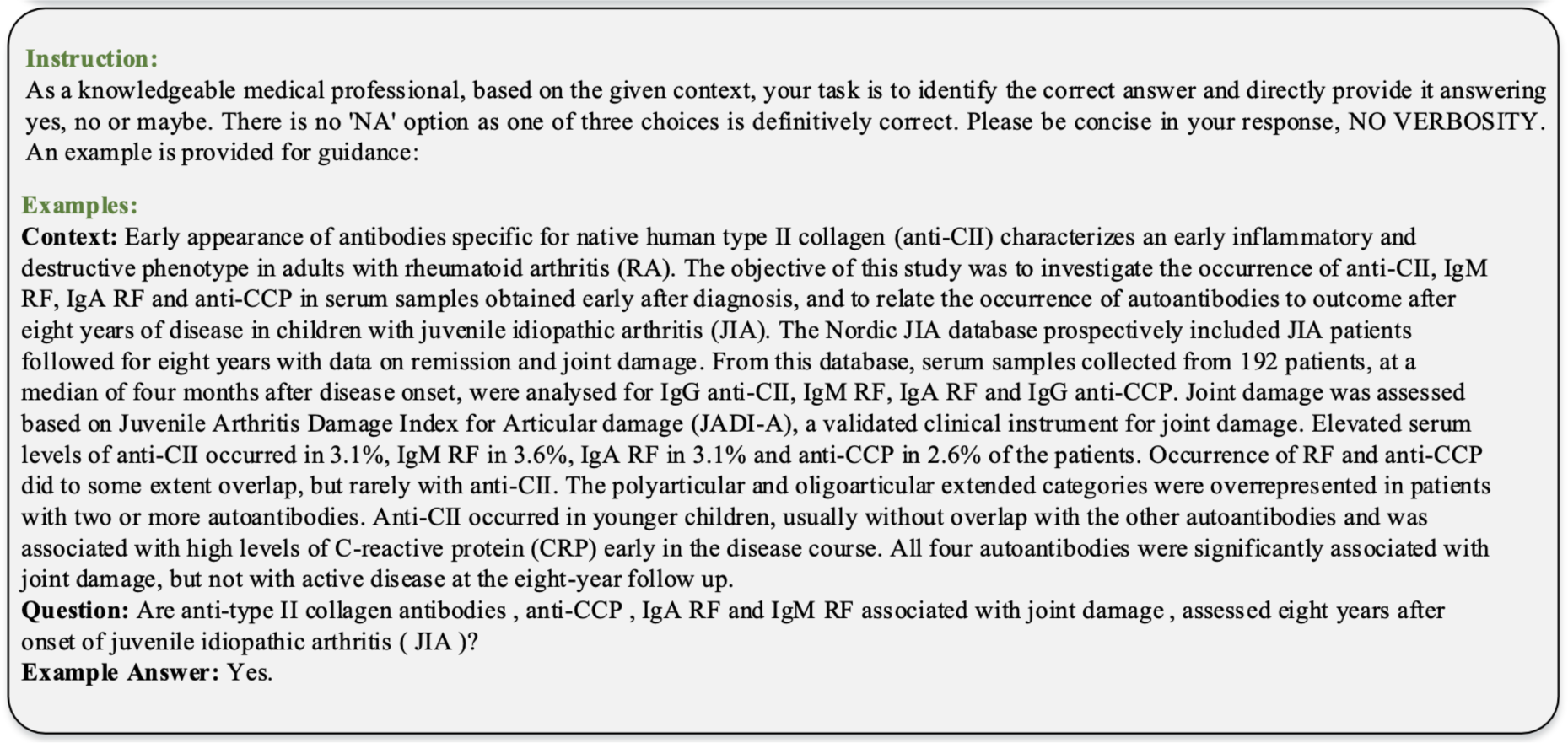

Prompt for MedQA-USMLE dataset, English language:

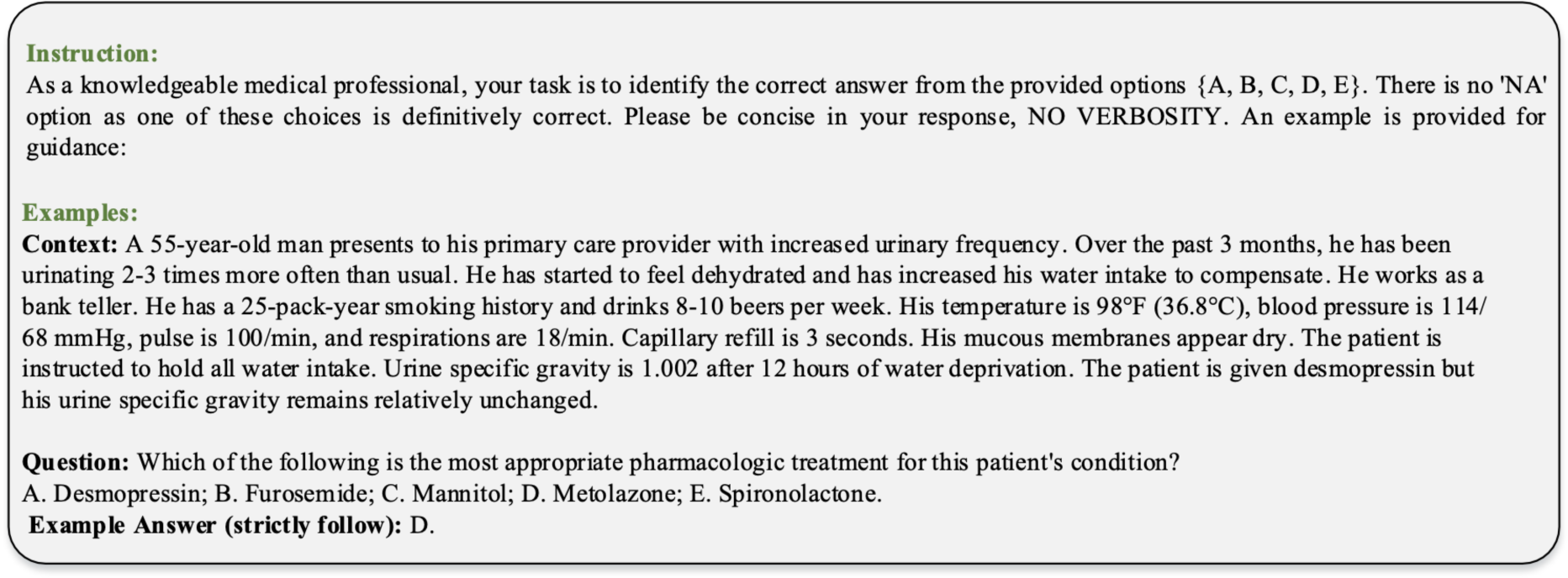

